# Temporal Trends in Stroke Management and Outcomes between 2011 and 2020: Results from a Nationwide Multicenter Registry

**DOI:** 10.1101/2024.02.29.24303345

**Authors:** Tai Hwan Park, Keun-Sik Hong, Yong-Jin Cho, Wi-Sun Ryu, Dong-Eog Kim, Man-Seok Park, Kang-Ho Choi, Joon-Tae Kim, Jihoon Kang, Beom-Joon Kim, Moon-Ku Han, Jun Lee, Jae-Kwan Cha, Dae-Hyun Kim, Jae Guk Kim, Soo Joo Lee, Jee-Hyun Kwon, Wook-Joo Kim, Dong-Ick Shin, Kyu Sun Yum, Sung Il Sohn, Jeong-Ho Hong, Jay Chol Choi, Byung-Chul Lee, Kyung-Ho Yu, Mi-Sun Oh, Jong-Moo Park, Kyusik Kang, Kyungbok Lee, Sang-Hwa Lee, Hae-Bong Jeong, Kwang-Yeol Park, Ji Sung Lee, Juneyoung Lee, Philip B. Gorelick, Hee-Joon Bae

**Affiliations:** Seoul Medical Center, Department of Neurology, Seoul, Korea; Ilsan Paik Hospital, Inje University, Department of Neurology, Goyang, Korea; Dongguk University Ilsan Hospital, Department of Neurology, Goyang, Korea; Chonnam National University Hospital, Department of Neurology, Gwangju, Korea; Seoul National University Bundang Hospital, Department of Neurology, Cerebrovascular Center, Seongnam, Korea; Yeungnam University Hospital, Department of Neurology, Daegu, Korea; Dong-A University Hospital, Department of Neurology, Busan, Korea; Eulji University Hospital, Eulji University, Department of Neurology, Daejeon, Korea; Ulsan University College of Medicine, Department of Neurology, Ulsan, Korea; Chungbuk National University Hospital, Department of Neurology, Cheongju, Korea; Keimyung University Dongsan Medical Center, Department of Neurology, Daegu, Korea; Jeju National University Hospital, Jeju National University School of Medicine, Department of Neurology, Jeju, Korea; Hallym University Sacred Heart Hospital, Department of Neurology, Anyang, Korea; Uijeongbu Eulji Medical center, Eulji University, Department of Neurology, Uijenongbu, Korea; Nowon Eulji Medical Center, Eulji University School of Medicine, Department of Neurology, Seoul, Korea; Soonchunhyang University Hospital, Department of Neurology, Seoul, Korea; Hallym University Chuncheon Sacred Heart Hospital, Department of Neurology, Gangwon-do, Korea; Chung-Ang University Hospital, Department of Neurology, Seoul, Korea; Asan Medical Center, Clinical Research Center, Seoul, Korea; Korea University College of Medicine, Department of Biostatistics, Seoul, Korea; Northwestern University Feinberg School of Medicine, Davee Department of Neurology, Chicago, IL, USA

**Author notes:** **Corresponding author:** Hee-Joon Bae, MD, PhD, FAHA, Department of Neurology, Seoul National University College of Medicine, Seoul National University Bundang Hospital, 82 Gumi-ro 173 Beon-gil, Bundang-gu, Seongnam-si, Gyeonggi-do 13620, Republic of Korea.

## Abstract

**Background:** There have been significant advancements in the treatment of ischemic stroke including stent retrievers for endovascular thrombectomy, new oral anticoagulants for atrial fibrillation, dual antiplatelet therapy for stroke prevention, and statins for atherosclerotic stroke. This study aims to evaluate temporal trends of these treatments and related clinical outcomes through a decade-long trend analysis, utilizing data from a comprehensive, national, multicenter stroke registry. We also seek to identify areas in need of improvement.

**Methods:** This analysis involved patients with ischemic stroke or transient ischemic attack registered prospectively in the Clinical Research Center for Stroke-Korea-National Institute of Health (CRCS-K-NIH) registry between 2011 and 2020. We examined temporal trends in risk factors, etiologic subtypes, acute management strategies, and outcomes for up to one year post-stroke. Generalized linear mixed models were employed to account for center clustering.

**Results:** Among 77,662 patients over 10 years, the average age increased by 2.2 years in men and 2.4 years in women. Notably, in-hospital neurological deterioration, 3-month and 1-year mortality, and cumulative incidence of recurrent stroke within one year showed significant decreases over time after adjustments for age, sex, and initial stroke severity (*P_trend_’s* < 0.01). However, functional outcomes at 3 months and 1 year remained unchanged. The use of endovascular thrombectomy increased from 5.4% in 2011 to 10.6% in 2020. There was also an increase in the prescription of anticoagulants for atrial fibrillation, dual antiplatelet therapy, statins, and stroke unit care. Contrarily, the rate of intravenous thrombolysis showed a slight decline.

**Conclusions:** This study points to a reduction in mortality and risk of recurrent stroke over the past decade, paralleling enhancement in acute and preventive stroke management. Nevertheless, the decline in use of intravenous thrombolysis and the stagnation of functional outcomes may signal the need for further investigation to identify underlying causes of these trends and counterstrategies to mitigate risks.

## Introduction

Over the last decade, ischemic stroke treatment has seen significant advancements with the introduction of stent retrievers for endovascular thrombectomy (EVT),^1^ novel oral anticoagulants (NOACs) for atrial fibrillation,^2–4^ dual antiplatelet therapy (DAPT) for minor strokes or high-risk transient ischemic attacks (TIAs),^5,6^ and high-dose statins for atherosclerotic stroke prevention.^7,8^ While these treatments have been successfully integrated into clinical practice, their comprehensive impact on clinical outcomes warrants further investigation.

Evaluating these treatments’ effectiveness requires examining both of stroke recurrence and functional outcomes, as their impacts may vary based on the outcome measured. For example, appropriate antithrombotic selection can reduce stroke recurrence, ^11^ and wider use of recanalization therapies and strategies preventing neurological deterioration during hospitalization may enhance functional outcomes.^1,12^

Despite a reported decline in stroke mortality and case fatality rates,^13–16^ insights into non-mortality stroke outcomes remain limited. Previous studies, like a German population-based study^17^ on recurrent stroke risk over 20 years and a comprehensive Japanese study of ischemic stroke cases^18^ reporting improving functional outcomes at discharge between 2000 and 2019, have not fully addressed long-term outcomes or the nuances of early treatment and stroke characteristics.

The Clinical Research Collaboration for Stroke in Korea-National Institute of Health (CRCS-K-NIH) registry, initiated in 2008, offers a rich dataset on stroke characteristics, treatments, and outcomes.^19^ Utilizing this registry database, we aim to outline trends in stroke outcomes, including functional outcomes at 3 months and 1 year, major vascular events, and mortality within the first year. It also seeks to assess trends in acute stroke treatments from 2011 to 2020, aiming to pinpoint areas for quality enhancement.

## Methods

### Study Design and Participants

This trend analysis focused on acute ischemic stroke (AIS) or TIA patients admitted from January 1, 2011, to December 31, 2020, identified within the CRCS-K-NIH registry.^19,20^ The registry includes demographics, clinical presentation, risk factors, treatments, and quality indicators, collected via a web-based system following predefined protocols.

### Outcome Measures

We analyzed in-hospital neurological deterioration (ND), 3-month and 1-year functional outcomes, and major vascular events and mortality within the first year, assessed through structured interviews by trained nurse coordinators or physicians during follow-up.

### Data Quality and Fidelity

Data integrity was maintained via monthly downloads, verification, steering committee reviews, and audits. Further details on quality and fidelity of data are available on the CRCS-K website (http://crcs-k.strokedb.or.kr/eng/).^19,20^

### Study variables

Evaluated variables included traditional stroke risk factors, pre-stroke medications, stroke severity (NIH Stroke Scale [NIHSS]), stroke onset, onset-to-arrival (OTA) time, and stroke etiological subtypes. Diagnostic assessments covered diffusion-weighted imaging (DWI), perfusion-weighted imaging (PWI), CT/MR angiography, echocardiography, and 24-hour Holter monitoring. Treatment variables encompassed intravenous thrombolysis (IVT) and EVT use, door-to-needle (DTN) and door-to-puncture (DTP) times, and discharge medications. Outcomes measured included ND, symptomatic hemorrhagic transformation, hospital stay length, recurrent stroke, modified Rankin Scale (mRS) scores, and mortality. Detailed descriptions of the registry’s design and data collection are documented in prior publications.^19,20^

### Ethics statement

Institutional review boards (IRBs) at all involved centers approved the collection of clinical information in the CRCS-K-NIH registry to monitor and improve the quality and outcomes of stroke care, granting a waiver of informed consent due to data anonymization and minimal participant risk. The IRBs also approved this research’s specific use of the registry database.

### Statistical analysis

Baseline characteristics, acute treatments, and outcomes were analyzed by calendar year, focusing on the proportions of participants with mild (NIHSS scores <4) and severe strokes (NIHSS scores ≥15), OTA time, and the promptness of IVT and EVT administration. We standardized the treatment of missing values and outliers, especially for DTN and DTP times, and length of hospital stay. The details are presented in supplemental material.

Linear regression was applied to continuous variables, and the Cochran–Armitage test to binary variables, to identify temporal trends. We utilized generalized linear mixed models with a random center effect to control for variations in age, sex, and center in examining baseline characteristics, diagnostic evaluations, and discharge medications. The initial NIHSS score was also adjusted in analyses related to acute treatment and outcomes. The usage rate of DAPT at discharge was specifically calculated for patients with non-cardioembolic minor stroke or high-risk TIA.

Visual representations of recanalization therapy trends were depicted in bar graphs for three epochs: 2011–2014, 2015–2017, and 2018–2020, to underscore shifts, particularly post-mid-2010s. Statistical significance was set at a 5% level, with all P values being two-sided. SAS version 9.4 (SAS Institute, Inc., Cary, NC) was employed for statistical analysis.

### Data availability

The data analyzed in this study are available from the corresponding author upon reasonable request.

## Results

From 2011 to 2020, 77,662 AIS or TIA patients were enrolled in the CRCS-K-NIH registry, with TIA cases constituting 9.3% (n=7,212).

### Baseline characteristics

The decade saw an increase in the average age of patients (2.2 years for men, 2.4 years for women) and a 1.8% rise in male patient proportion (Table 1). Despite an older stroke demographic, major modifiable risk factors remained stable. Current smoking decreased by 2.6%, and new dyslipidemia diagnoses fell by 28.8%. Oral anticoagulant and statin use before stroke consistently rose by 27.7% and 10.1%, respectively. Stroke severity at presentation decreased, with the mean initial NIHSS score dropping from 6.0 to 5.4. Mild stroke (NIHSS <4) instances rose, whereas severe strokes (NIHSS ≥15) declined. Emergency room admissions increased by 2.9%, with no significant change in OTA times. Large artery atherosclerosis (LAA) was the leading ischemic stroke subtype, with slight shifts in subtype proportions over time.

**Table 1.** Secular Trends in Baseline Characteristics.

| Year of admission | Total | 2011 | 2012 | 2013 | 2014 | 2015 | 2016 | 2017 | 2018 | 2019 | 2020 | $P_{trend}$ | Adjusted $P_{trend}^*$ | $\beta$ |
| --- | --- | --- | --- | --- | --- | --- | --- | --- | --- | --- | --- | --- | --- | --- |
| Number of patients | 77,662 | 5,660 | 5,978 | 6,900 | 7,458 | 7,670 | 8,515 | 8,864 | 8,674 | 9,513 | 8,430 |  |  |  |
| Male, % | 58.4 | 58.0 | 57.4 | 58.3 | 58.1 | 57.3 | 58.2 | 59.7 | 57.6 | 59.3 | 59.8 | 0.04 | <.01 | + |
| Age, y, mean (SD) |  |  |  |  |  |  |  |  |  |  |  |  |  |  |
| Men | 65.6 (12.7) | 64.7 (12.7) | 65.2 (12.6) | 64.5 (12.8) | 65.1 (13.0) | 65.3 (12.4) | 65.3 (12.6) | 65.8 (12.7) | 65.8 (12.8) | 66.4 (12.8) | 66.9 (12.4) | <.01 | <.01 | + |
| Women | 71.6 (12.8) | 70.5 (12.4) | 70.7 (12.6) | 70.7 (12.8) | 71.0 (12.6) | 71.2 (12.5) | 71.4 (12.9) | 71.9 (12.9) | 72.5 (12.8) | 72.4 (13.3) | 72.9 (13.2) | <.01 | <.01 | + |
| Risk factors, % |  |  |  |  |  |  |  |  |  |  |  |  |  |  |
| Hypertension | 65.9 | 69.3 | 67.6 | 66.1 | 67.0 | 62.8 | 64.3 | 64.0 | 66.4 | 65.5 | 67.5 | <.01 | <.01 | - |
| Newly diagnosed | 9.0 | 11.2 | 9.4 | 9.7 | 7.7 | 7.3 | 8.7 | 8.7 | 9.4 | 9 | 9.7 | 0.12 | 0.12 | + |
| Diabetes | 32.4 | 33.4 | 32.6 | 32.2 | 32.5 | 31.2 | 31.6 | 31.5 | 31.9 | 32.8 | 34.6 | 0.18 | 0.87 | + |
| Newly diagnosed | 12.6 | 13.8 | 12.1 | 14.4 | 10.9 | 10.3 | 12.1 | 13.3 | 12.3 | 12.4 | 14.1 | 0.28 | 0.54 | + |
| Dyslipidemia | 30.7 | 39.1 | 32.0 | 27.9 | 26.0 | 25.6 | 30.8 | 30.5 | 28.6 | 32.8 | 34.6 | 0.13 | <.01 | + |
| Newly diagnosed | 35.2 | 56.5 | 42.8 | 36.1 | 33 | 35.1 | 36.6 | 32.5 | 28.3 | 30.1 | 27.7 | <.01 | <.01 | - |
| Atrial fibrillation | 20.0 | 19.7 | 20.8 | 18.9 | 21.1 | 21.5 | 19.5 | 18.9 | 20.1 | 19.6 | 20.1 | 0.25 | <.01 | - |
| Newly diagnosed | 45.9 | 46.4 | 38.5 | 40.9 | 46.5 | 46.7 | 45.6 | 47.7 | 45.9 | 48.4 | 49.6 | <.01 | <.01 | + |
| History of stroke | 20.7 | 21.3 | 20.4 | 20.8 | 21.1 | 20.1 | 20.2 | 19.9 | 20.7 | 21.2 | 21.7 | 0.31 | 0.15 | + |
| Current smoking | 23.0 | 25.0 | 25.4 | 25.7 | 24.0 | 20.5 | 22.9 | 23 | 21.2 | 21.9 | 22.4 | <.01 | <.01 | - |
| Admitted via ER, % | 89.1 | 87.7 | 86.8 | 87.1 | 87.6 | 87.6 | 88.5 | 90 | 90.6 | 92.2 | 90.6 | <.01 | <.01 | + |
| Unclear onset, % | 40.9 | 39.1 | 48.7 | 48.2 | 40.4 | 42.3 | 37.3 | 38.8 | 40.8 | 41.4 | 35.6 | <.01 | <.01 | - |
| From FAT to arrival, h, median (IQR) | 7.8 (2.2–34.2) | 8.0 (2.3–33.2) | 7.7 (2.1–34.4) | 7.1 (2.0–30.6) | 7.6 (2.0–34.6) | 6.8 (1.8–31.1) | 6.8 (1.9–29.5) | 6.8 (1.7–29.9) | 7.2 (1.8–31.9) | 7.8 (1.9–32.7) | 7.3 (1.9–32.0) | 0.27 | 0.052 | - |
| NIHSS at presentation |  |  |  |  |  |  |  |  |  |  |  |  |  |  |
| Mean (SD) | 5.3 (6.1) | 5.4 (6.0) | 5.5 (6.2) | 5.5 (6.2) | 5.7 (6.5) | 5.4 (6.3) | 5.3 (6.1) | 5.2 (5.9) | 5.3 (6.1) | 5.0 (5.9) | 5.1 (5.8) | <.01 | <.01 | - |
| NIHSS<4, % | 49.7 | 47.8 | 47.9 | 48.7 | 47.6 | 49.6 | 49.6 | 49.3 | 49.6 | 53.0 | 52.1 | <.01 | <.01 | + |
| NIHSS≥15, % | 11.9 | 12.4 | 12.3 | 12.7 | 13.7 | 12.8 | 12.2 | 11.1 | 11.8 | 10.8 | 10.4 | <.01 | <.01 | - |
| Pre-stroke, % |  |  |  |  |  |  |  |  |  |  |  |  |  |  |
| Antiplatelets | 28.7 | 30.7 | 29.4 | 29.1 | 30.2 | 30.2 | 26.6 | 26.9 | 28.2 | 28.1 | 29.0 | <.01 | <.01 | - |
| Oral Anticoagulants in atrial fibrillation | 41.1 | 28.4 | 25.2 | 29.0 | 36.1 | 38.3 | 45.3 | 43.6 | 48.3 | 51.4 | 56.1 | <.01 | <.01 | + |
| Statin | 20.8 | 15.1 | 17.7 | 17.4 | 19.1 | 20.3 | 20.0 | 21.0 | 22.7 | 25.0 | 25.2 | <.01 | <.01 | + |
| TOAST subtypes, %<sup>**</sup> |  |  |  |  |  |  |  |  |  |  |  |  |  |  |
| LAA | 35.0 | 37.5 | 38.7 | 36.6 | 33.9 | 33 | 35.4 | 34 | 34.2 | 33.2 | 35.6 | <.01 | <.01 | - |
| SVO | 17.7 | 17.1 | 15.7 | 17.1 | 16.7 | 17.7 | 17.7 | 19.6 | 19.1 | 18 | 17.5 | <.01 | 0.01 | + |
| CE | 21.2 | 22.0 | 22.2 | 21.4 | 23.1 | 23.3 | 21.0 | 20.1 | 21.2 | 20.1 | 19.2 | <.01 | <.01 | - |
| OE | 3.5 | 2.1 | 2.4 | 2.6 | 3.0 | 3.2 | 3.4 | 3.8 | 4.5 | 4.3 | 4.5 | <.01 | <.01 | + |
| UE (2 or more) | 4.4 | 4.8 | 4.4 | 4.3 | 4.2 | 4.8 | 4.6 | 4.7 | 3.8 | 4.2 | 4.4 | 0.29 | 0.01 | - |
| UE (negative) | 9.0 | 7.5 | 6.5 | 7.7 | 7.9 | 8.7 | 9.4 | 9.9 | 9.0 | 10.6 | 11.2 | <.01 | <.01 | + |
| UE (incomplete) | 9.1 | 9.0 | 10.0 | 10.4 | 11.1 | 9.3 | 8.6 | 8.0 | 8.2 | 9.6 | 7.5 | <.01 | <.01 | - |

ER, emergency room; FAT, first abnormal time; IQR, interquartile range; NIHSS, National Institutes of Health Stroke Scale; TOAST, Trial of ORG 10172 in Acute Stroke Treatment; LAA, large artery atherosclerosis; SVO, small vessel occlusion; CE, cardioembolism; OE, other etiology; UE, undetermined etiology.
The denominator of the newly diagnosed risk factor ratio is the frequency of risk factors.
\* Adjusted for age and sex with random effects for the center.
\*\* Among 70,450 patients excluding TIA
The $\beta$ coefficient is indicated as + or - to indicate the direction of the slope and, if the directions of the univariate analysis and the multivariate analysis are different, they are separated with “/”.

### Stroke management

Among 70,450 AIS patients, EVT usage increased by 5.2%, while IVT saw a 2% decline (Table 2). These trends were consistent across different patient groups and time frames, notably among older patients and those with cardioembolic (CE) subtype or pre-stroke oral anticoagulant (OAC) use (Figure). DTN times for IVT remained stable, but DTP times for EVT improved significantly, with a 6.1% increase in cases meeting the ≤60 min target. Increased utilization of stroke unit care, non-invasive cardiac evaluations, and prescriptions for DAPT, NOACs, and statins at discharge was observed. DAPT use notably doubled in patients with non-CE minor stroke or high-risk TIA, and statin in LAA and OAC use in atrial fibrillation increased, with NOACs surpassing warfarin since 2015.

**Table 2.**
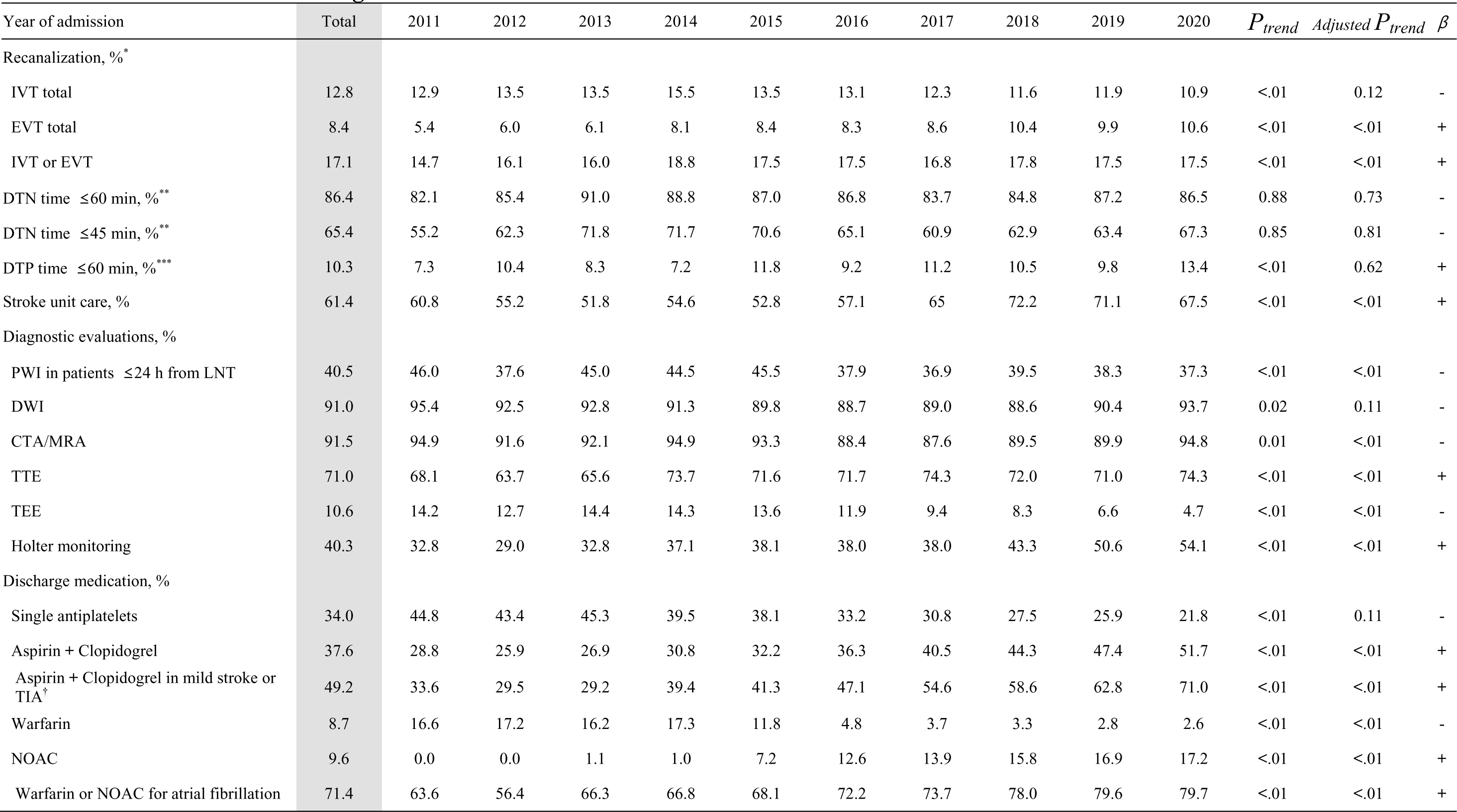
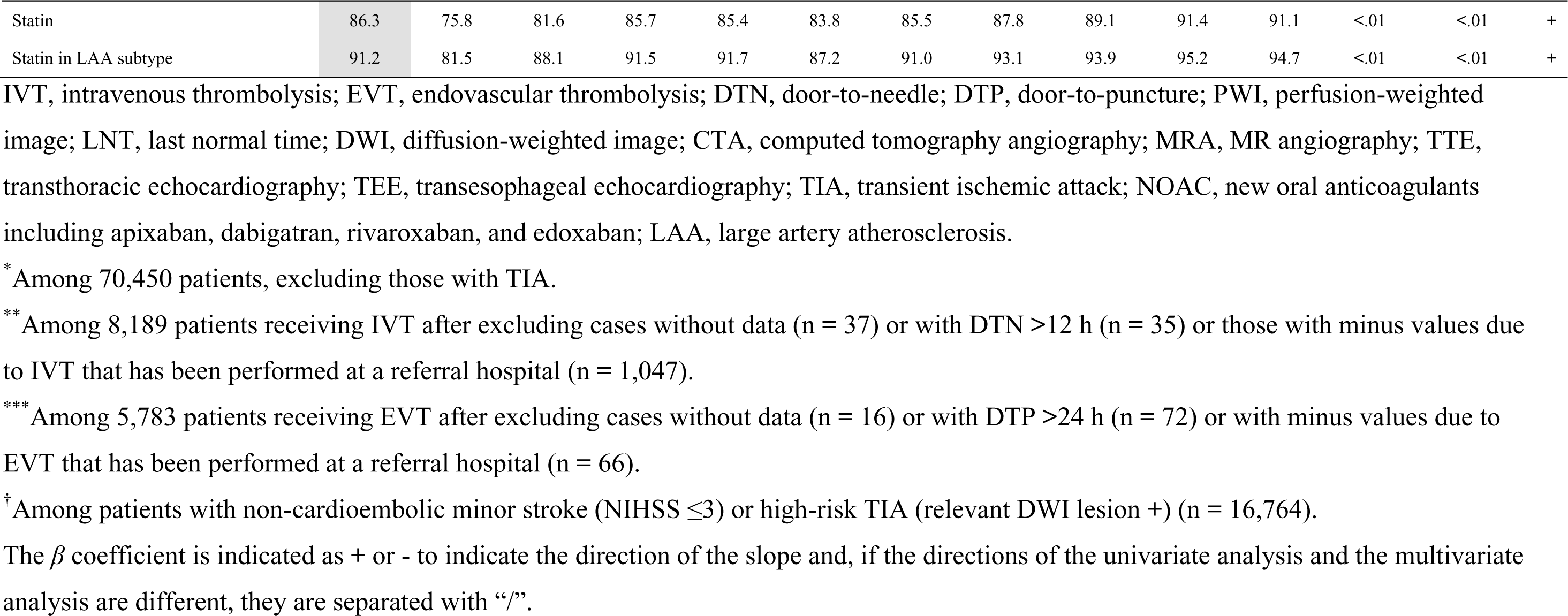
Secular Trends in Managements.

**Figure.**
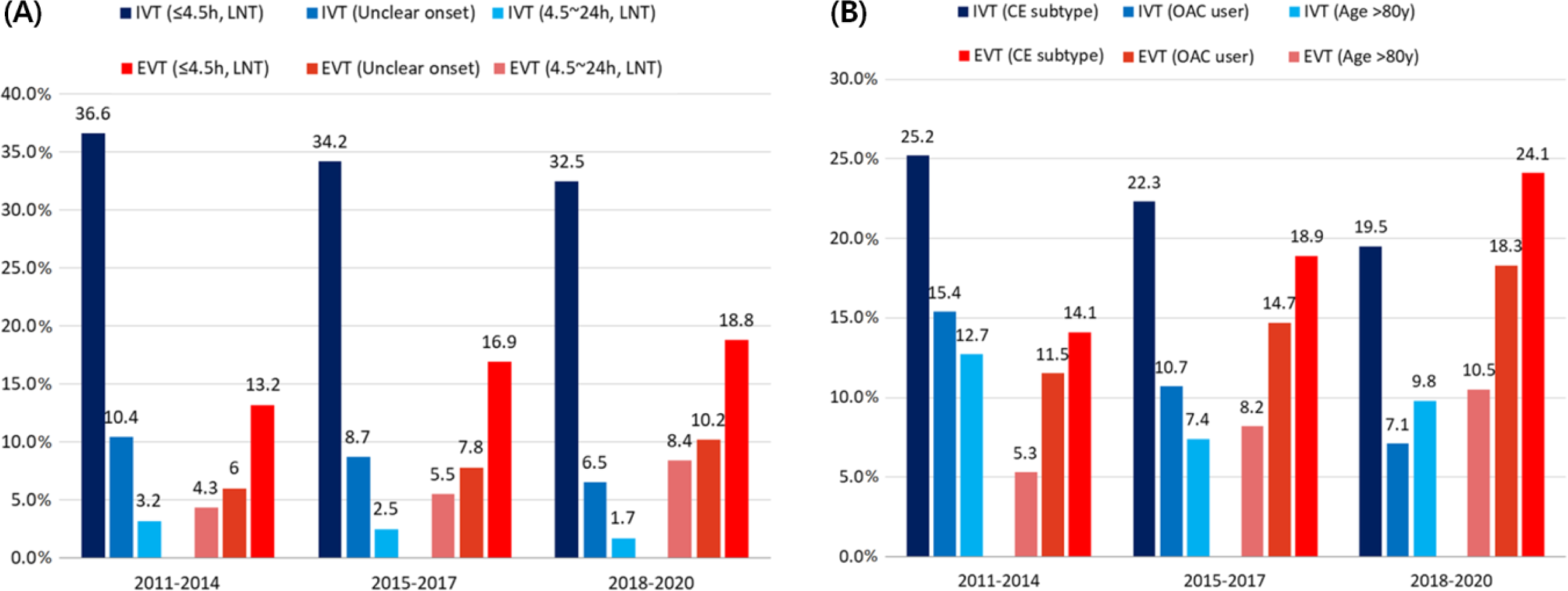
Changes of Recanalization Therapy Rates according to Hospital Arrival Time (A) and Various Clinical Features (B) During the study period, the IVT rate decreased in patients with various clinical conditions, while the EVT rate increased. CE indicates cardioembolism; EVT, endovascular thrombectomy; IVT, intravenous thrombolysis; LNT, last normal time; OAC, oral anticoagulants.

### Outcomes

ND during hospitalization occurred in average 10.4% of patients with AIS or TIA, showing a significant ten-year decline (crude and adjusted *P_trend_*’s < 0.01; Table 3). Conversely, symptomatic hemorrhagic transformation rates post-recanalization therapy remained stable at 3.0%. Length of hospital stay has decreased significantly.

**Table 3.**
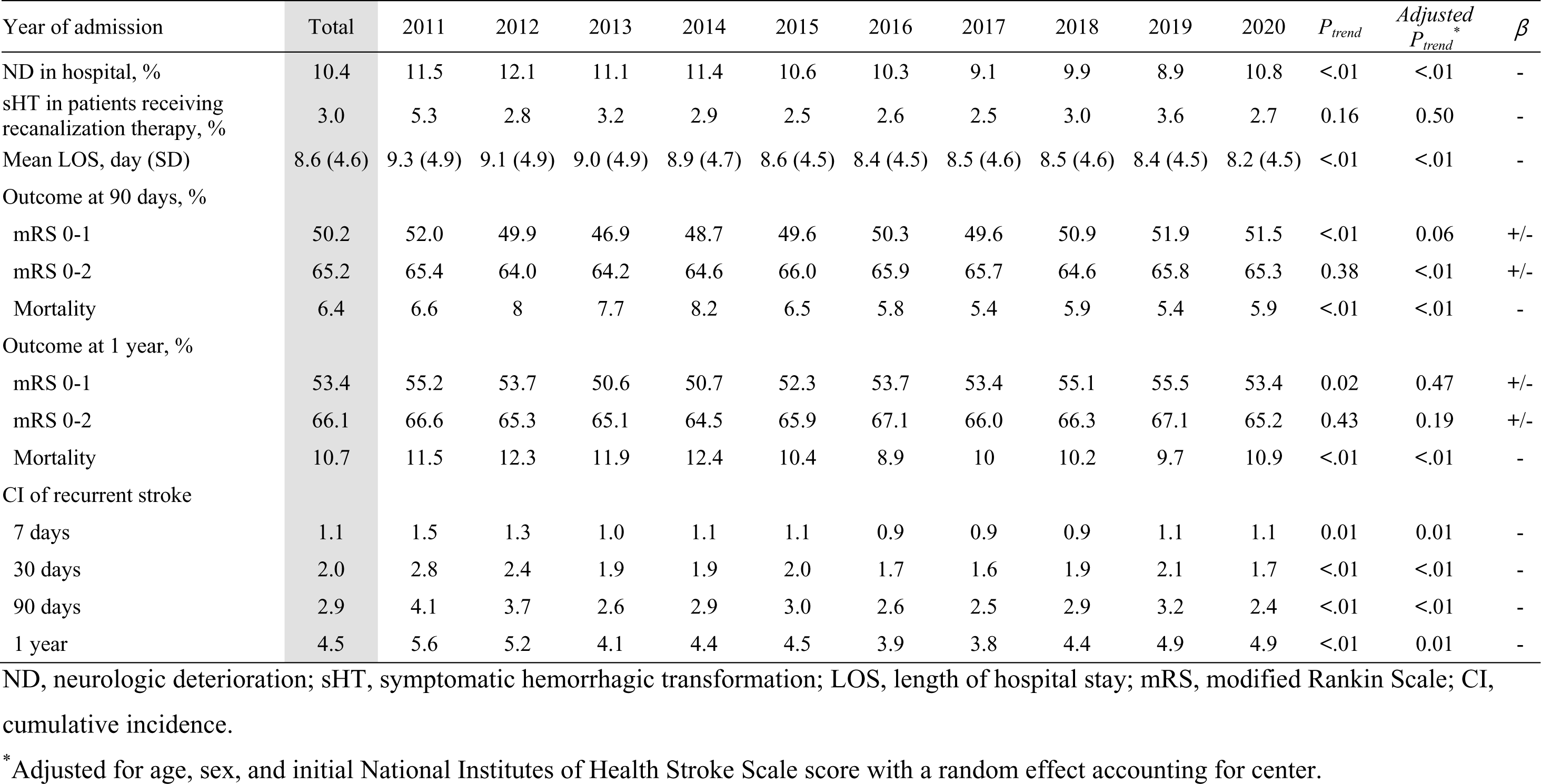
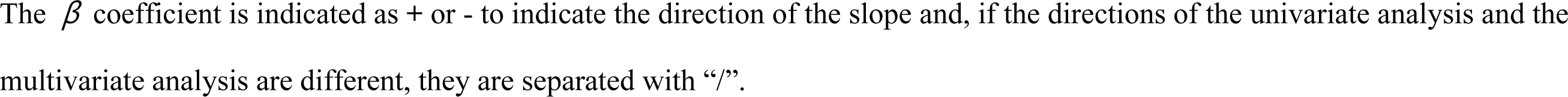
Secular Trends in Stroke Outcome.

| Year of admission | Total | 2011 | 2012 | 2013 | 2014 | 2015 | 2016 | 2017 | 2018 | 2019 | 2020 | $P_{trend}$ | *Adjusted*<br>$P_{trend}^*$ | $\beta$ |
| --- | --- | --- | --- | --- | --- | --- | --- | --- | --- | --- | --- | --- | --- | --- |
| ND in hospital, % | 10.4 | 11.5 | 12.1 | 11.1 | 11.4 | 10.6 | 10.3 | 9.1 | 9.9 | 8.9 | 10.8 | <.01 | <.01 | - |
| sHT in patients receiving recanalization therapy, % | 3.0 | 5.3 | 2.8 | 3.2 | 2.9 | 2.5 | 2.6 | 2.5 | 3.0 | 3.6 | 2.7 | 0.16 | 0.50 | - |
| Mean LOS, day (SD) | 8.6 (4.6) | 9.3 (4.9) | 9.1 (4.9) | 9.0 (4.9) | 8.9 (4.7) | 8.6 (4.5) | 8.4 (4.5) | 8.5 (4.6) | 8.5 (4.6) | 8.4 (4.5) | 8.2 (4.5) | <.01 | <.01 | - |
| Outcome at 90 days, % |  |  |  |  |  |  |  |  |  |  |  |  |  |  |
| mRS 0-1 | 50.2 | 52.0 | 49.9 | 46.9 | 48.7 | 49.6 | 50.3 | 49.6 | 50.9 | 51.9 | 51.5 | <.01 | 0.06 | +/- |
| mRS 0-2 | 65.2 | 65.4 | 64.0 | 64.2 | 64.6 | 66.0 | 65.9 | 65.7 | 64.6 | 65.8 | 65.3 | 0.38 | <.01 | +/- |
| Mortality | 6.4 | 6.6 | 8 | 7.7 | 8.2 | 6.5 | 5.8 | 5.4 | 5.9 | 5.4 | 5.9 | <.01 | <.01 | - |
| Outcome at 1 year, % |  |  |  |  |  |  |  |  |  |  |  |  |  |  |
| mRS 0-1 | 53.4 | 55.2 | 53.7 | 50.6 | 50.7 | 52.3 | 53.7 | 53.4 | 55.1 | 55.5 | 53.4 | 0.02 | 0.47 | +/- |
| mRS 0-2 | 66.1 | 66.6 | 65.3 | 65.1 | 64.5 | 65.9 | 67.1 | 66.0 | 66.3 | 67.1 | 65.2 | 0.43 | 0.19 | +/- |
| Mortality | 10.7 | 11.5 | 12.3 | 11.9 | 12.4 | 10.4 | 8.9 | 10 | 10.2 | 9.7 | 10.9 | <.01 | <.01 | - |
| CI of recurrent stroke |  |  |  |  |  |  |  |  |  |  |  |  |  |  |
| 7 days | 1.1 | 1.5 | 1.3 | 1.0 | 1.1 | 1.1 | 0.9 | 0.9 | 0.9 | 1.1 | 1.1 | 0.01 | 0.01 | - |
| 30 days | 2.0 | 2.8 | 2.4 | 1.9 | 1.9 | 2.0 | 1.7 | 1.6 | 1.9 | 2.1 | 1.7 | <.01 | <.01 | - |
| 90 days | 2.9 | 4.1 | 3.7 | 2.6 | 2.9 | 3.0 | 2.6 | 2.5 | 2.9 | 3.2 | 2.4 | <.01 | <.01 | - |
| 1 year | 4.5 | 5.6 | 5.2 | 4.1 | 4.4 | 4.5 | 3.9 | 3.8 | 4.4 | 4.9 | 4.9 | <.01 | 0.01 | - |

ND, neurologic deterioration; sHT, symptomatic hemorrhagic transformation; LOS, length of hospital stay; mRS, modified Rankin Scale; CI, cumulative incidence.
\* Adjusted for age, sex, and initial National Institutes of Health Stroke Scale score with a random effect accounting for center.
The $\beta$ coefficient is indicated as + or - to indicate the direction of the slope and, if the directions of the univariate analysis and the multivariate analysis are different, they are separated with “/”.

Follow-up data revealed that 3-month outcomes were documented for 96.2% of participants, and one-year outcomes for 92.1%. There was a marked reduction in mortality rates at both 3 and 12 months (Table 3), peaking in 2014 (8.2% at 3 months and 12.4% at 9 months) before showing a downward trend. Additionally, we observed a significant decrease in the cumulative incidence of recurrent stroke up to one year (crude and adjusted *P_trend_*’s < 0.05). This decline in stroke recurrence varied across etiologic subtypes, with LAA and CE subtypes experiencing notable reductions, whereas the small vessel occlusion subtype did not (eTable 1).

An initial analysis indicated an increase in the proportion of patients achieving a mRS score of 0–1 at 3 and 12 months. However, this trend did not persist after adjusting for initial stroke severity (Table 3 and eTable 2). No significant improvement was observed in the proportion of patients with an mRS of 0–2 over the decade.

## Discussion

In our analysis of 77,662 patients with AIS or TIA from the 2011-2020 time period in a nationwide multicenter stroke registry, we observed improvement in most stroke outcomes across an aging demographic. This period was characterized by the wide adoption of advanced treatments like EVT, NOAC, DAPT, statin, and stroke unit care. Despite these advancements, we observed a stagnation in functional outcomes, and a decline in the utilization of IVT over the years.

Our study might be the first one to document changes in the occurrence of ND during hospitalization, a condition often linked to stroke progression or recurrence and associated with poorer outcomes.^12^ Additionally, the observed decline in stroke recurrence, especially in the LAA and CE subtypes, is also a notable finding. The reductions in ND and stroke recurrence could be explained at least in part by increased use of stroke unit care,^21^ DAPT,^5,6^ NOAC,^2–4,22^ and statin.^7,8^ However, direct evidence of these treatments preventing ND is yet to be established. Enhanced diagnostic approaches, including more frequent use of DWI, angiographic, and cardiac evaluations, also may have contributed to these improvements through better identification and leading to treatment of specific underlying causes of stroke.

The observed decline in ischemic stroke mortality in recent decades^13–15^ can be attributed to better control of vascular risk factors^23^ and advancements in in-hospital management.^24^ Interestingly, this decline became more pronounced following 2015, aligning with the seminal EVT trials,^25–27^ with our data indicating a peak in mortality in 2014 followed by a notable decrease from 2015 onwards (Table 3). Additionally, the observed reduction in stroke severity may have contributed to improved outcomes. However, our analysis, adjusted for stroke severity, indicates that outcomes have improved independently of stroke severity, suggesting other contributing factors to these positive trends.

Contrary to other studies indicating a rise in major vascular risk factors,^14,16,28^ our study found an unexpected decrease in the prevalence of hypertension and atrial fibrillation, despite an older population (Table 1). In Korea, hypertension management has improved considerably, leading to a reduced prevalence of hypertension in the general population from 1998 to 2018.^29^ Additionally, the increased use of oral anticoagulants for atrial fibrillation, particularly since the introduction of NOAC reimbursement in South Korea for high-risk patients in 2015, may explain the declining prevalence in AIS patients.^9^ The rise in newly diagnosed atrial fibrillation cases could be attributed to enhanced primary prevention and increased cardiac evaluation. However, the stable proportion of most newly diagnosed risk factors during hospitalization over the decade suggests that primary prevention efforts still leave room for improvement.

The observed reductions in LAA and CE subtypes may also be linked to improved preventive strategies. The decline in LAA, mirroring trends seen in Western countries,^30^ could be attributed to increased statin use. Similarly, the reduction in CE might be due to the expanded use of NOACs in patients with atrial fibrillation. The decrease in initial stroke severity observed in our study may also result from prestroke statin^31^ or NOAC administration.^32^ The increase in strokes of unknown etiology highlights the potential need for broader application of advanced diagnostic tools, such as implantable loop recorders, to enhance identification of stroke etiology.

Surprisingly, we observed no improvement in functional outcomes during our study period, contrary to expectations based on international results. A recent Japanese study reported improved functional outcomes with increase in use of acute reperfusion therapy in AIS patients over two decades.^18^ However, in our study, the IVT administration rate decreased post-2014, unlike in other countries where it has steadily risen in recent years.^18,33,34^ Notably, functional outcomes seemed to improve in IVT-treated patients during our study period (eTable 3). This suggests that the declining IVT rate might be a contributing factor to the lack of improvement in overall functional outcomes, despite an increase in EVT rates. This finding highlights a potential area for further investigation and improvement in stroke care strategies.

The decline in IVT rates in our study can be attributed to several factors: a higher proportion of patients with mild strokes, increased pre-stroke use of oral anticoagulants, and more comorbidities related to an aging population. The simultaneous decrease in IVT and increase in EVT, even among patients arriving within 4.5 hours of stroke or TIA onset, suggests that a rise in IVT-ineligible patients may be driving the reduced IVT rate. This is supported by findings from the Swiss Stroke Registry, which reported a notably low IVT rate in ischemic stroke patients with atrial fibrillation on NOACs.^32^ This contrasts with recent research suggesting the safety of IVT in patients who have taken NOACs within 48 hours before stroke onset.^35^

The lack of improvement in OTA time highlights another potential area in need of improvement. Innovations in prehospital stroke systems of care are necessary, despite shortened DTN times and decreased ND after recanalization therapy (eTable 4, 5). Additionally, despite pivotal trials extending the recanalization therapy window,^36,37^ the use of perfusion imaging in patients arriving within 24 hours from the LNT and those with unclear onset (accounting for 41% of our study population) was lower than anticipated. This underutilization represents another potential area in need of novel strategies to improve stroke functional outcomes.

Our study’s strengths include a large sample size and consistent data collection using standardized protocols since 2008, encompassing a wide range of outcome variables up to one year post-stroke. This allowed for a comprehensive examination of trends in stroke outcomes and their key determinants, such as stroke severity, etiological subtypes, and acute treatments.

However, there are study limitations to consider. First, our participants may not fully represent the South Korean stroke population at large, as the CRCS-K-NIH registry primarily includes tertiary or training hospitals engaged in continuous quality improvement activities. Thus, the rapid adoption of advanced stroke treatments and associated outcomes may be more favorable in our study than in general hospitals. Nonetheless, the demographics and clinical features of our study participants align with those in a national stroke audit program covering over 200 acute-care hospitals in South Korea (eTable 6). Additionally, as our study population was predominantly Korean and all participating hospitals were in South Korea, ethnic and cultural differences might limit the generalizability of our findings to other settings. Second, the observational nature of this study precludes determining causality between treatment changes and outcomes. For example, a detailed analysis to explain the lack of improvement in functional outcomes is beyond the study’s scope, but warrants additional research to assess the impact of advanced treatments on functional recovery in real-world settings. Third, despite adjusting for age, sex, and stroke severity, residual confounding factors such as comorbidities, socioeconomic status, and medication adherence might have influenced the results.

In summary, our study reveals that in South Korea over the past decade, most clinical outcomes for patients with AIS or TIA have improved, with the notable exception of functional outcomes. During this period there also has been a steady increase in the use of treatments proven to be effective in acute stroke treatment and subsequent prevention.

However, our results highlight several challenges that need to be addressed to improve quality of stroke care. Additional efforts to develop prehospital stroke systems to reduce OTA time and increase the IVT rate promise to prove useful. Furthermore, research to investigate whether the observed trends, including decreased IVT rates and stagnation of functional outcomes, are unique to South Korea or represent broader trends in countries with older populations and represent an area worthy of risk mitigation related to the root causes of the findings.

## Supporting information

Supplemental eTable

