## Supplemental eTable for "Temporal Trends in Stroke Management and Outcomes between 2011 and 2020: Results from a Nationwide Multicenter Registry"

### Supplemental Methods

#### *Treatment of missing values or outliers*

Missing or outliers were dealt with in a pre-defined manner as follows: for subjects whose first-found abnormal time (FAT) to arrival time exceeds 7 days (n=2,847, 3.7%), the FAT to arrival time was substituted by the median value of the all study subjects; Door-to-needle (DTN) time was evaluated in 8,189 patients after excluding subjects without data (n=37) or with DTN >12hr (n=35) or minus values due to Intravenous thrombolysis (IVT) performed at referral hospital (n=1047); Door-to-Puncture (DTP) time was evaluated in 5,783 patients after excluding subjects without data (n=16) or with DTP time >24hr (n=72) or with minus values due to endovascular thrombectomy (EVT) performed at referral hospital (n=66); length-of-stay in hospital was evaluated in 72,278 patients with 2-26 days of hospitalization within one SD value.

**List of supplemental tables**

**eTable 1. Secular trends in In-hospital Neurologic Deterioration and Stroke Recurrence According to Etiologic Stroke Subtypes**

**eTable 2. Impact of Calendar Year on Functional Outcomes Post-Stroke**

**eTable 3. Three-Month Functional Outcomes in Patients receiving IVT or EVT**

**eTable 4. Secular Trends in Onset-To-Treatment Time among Patients Receiving IVT or EVT**

**eTable 5. Secular Trends in Incidence of Neurologic Deterioration During Hospitalization in Patients Receiving IVT or EVT**

**eTable 6. Comparisons of Patient Characteristics between the CRCS-K-NIH registry and National Stroke Audit Program**

**eTable 1. Secular trends in In-hospital Neurologic Deterioration and Stroke Recurrence According to Etiologic Stroke Subtypes**

| Year of admission | | Total | 2011-2014 | 2015-2017 | 2018-2020 | $P_{trend}$ | *$Adjusted\ P_{trend}$ | $\beta$ |
|---|---|---|---|---|---|---|---|---|
| ND in hospital, % | | | | | | | | |
| LAA | | 13.6 | 14.4 | 13.3 | 13.0 | 0.010 | 0.025 | - |
| CE | | 11.2 | 13.4 | 10.3 | 9.7 | <.001 | <.001 | - |
| SVO | | 8.2 | 8.2 | 8.3 | 8.1 | 0.657 | 0.551 | + |
| CI of recurrent stroke, % | | | | | | | | |
| LAA | 90 days | 3.5 | 3.9 | 3.3 | 3.3 | 0.002 | <.001 | - |
| | 1 year | 5.2 | 5.5 | 4.7 | 5.4 | 0.171 | 0.012 | - |
| CE | 90 days | 3.4 | 3.6 | 3.4 | 3.1 | 0.013 | 0.014 | - |
| | 1 year | 4.9 | 5.3 | 4.6 | 4.7 | 0.034 | 0.059 | - |
| SVO | 90 days | 1.3 | 1.5 | 1.3 | 1.3 | 0.355 | 0.754 | - |
| | 1 year | 2.3 | 2.2 | 2.2 | 2.6 | 0.394 | 0.676 | + |

CI indicates cumulative incidence; ND, neurologic deterioration; LAA, large artery atherosclerosis; SVO, small vessel occlusion; CE, cardioembolism.

*Adjusted for age, sex, initial NIHSS score

The $\beta$ coefficient was indicated as + or - to indicate the direction of the slope, and if the directions of the univariate analysis and the multivariate analysis were different, they were indicated by separating them with "/".

**eTable 2. Impact of Calendar Year on Functional Outcomes Post-Stroke**

| Outcome | | | | OR (95% CI) per 10 years | | |
|---|---|---|---|---|---|---|
| | | Total, N | mRS 0-1, N (%) | Unadjusted | Model 1 | Model 2 |
| mRS 0-1 | 3 months | 74,720 | 37,506 (50.2) | 1.051 (1.010-1.094) | 1.083 (1.020-1.149) | 0.942 (0.886-1.000) |
| | 1 year | 71,574 | 38,248 (53.4) | 1.062 (1.020-1.105) | 1.138 (1.062-1.219) | 0.970 (0.904-1.051) |

CI indicates confidence interval; mRS, modified Rankin Scale; OR, odds ratio.

Model 1 is adjusted for age, sex.

Model 2 is adjusted for age, sex, initial NIHSS score

**eTable 3. Three-Month Functional Outcomes in Patients Receiving IVT or EVT**

| | Treatment | Epoch | | | Total | $P_{trend}$ | *Adjusted $P_{trend}$ |
|---|---|---|---|---|---|---|---|
| | | 2011-2014 | 2015-2017 | 2018-2020 | | | |
| mRS 0-1, % | IVT (n=8968) | 34.0 | 40.2 | 43.0 | 38.8 | <.001 | 0.004 |
| | EVT (n=5752) | 25.5 | 28.9 | 29.4 | 28.2 | 0.063 | 0.052 |
| mRS 0-2, % | IVT (n=8968) | 49.4 | 56.8 | 57.0 | 54.2 | <.001 | 0.077 |
| | EVT (n=5752) | 39.3 | 42.8 | 41.7 | 41.4 | 0.426 | 0.400 |

EVT indicates endovascular thrombectomy; IVT, intravenous thrombolysis.

*Adjusted for age, sex, initial NIHSS score

**eTable 4. Secular Trends in Onset-To-Treatment Time among Patients Receiving IVT or EVT**

| Treatment | Time | Epoch | | | $P_{trend}$ | *Adjusted $P_{trend}$ |
|---|---|---|---|---|---|---|
| | | 2011-2014 | 2015-2017 | 2018-2020 | | |
| IVT (n=8189)** | OTN, median min. (IQR) | 112 (81-160) | 108 (76-160) | 103 (75-153) | 0.060 | 0.078 |
| EVT (n=5783)*** | OTP, median min. (IQR) | 215 (158-315) | 195 (143-272) | 198 (145-308) | 0.234 | 0.592 |

EVT indicates endovascular thrombectomy; IVT, intravenous thrombolysis; OTN, onset to needle time; OTP, onset to puncture time.

*Adjusted for age, sex, initial NIHSS score

**Among 8,189 patients receiving IV rtPA after excluding cases without data (n=37) or with DTN>12hr (n=35) or those with minus values due to IVT that has been performed refered hospital (n=1047).

***Among 5,783 patients receiving EVT after excluding cases without data (n=16) or with DTP>24hr (n=72) or with minus values due to EVT that has been performed refered hospital (n=66).

**eTable 5. Secular Trends in Incidence of Neurologic Deterioration During Hospitalization in Patients Receiving IVT or EVT**

| | Treatment | Epoch | | | Total | $P_{trend}$ | *Adjusted $P_{trend}$ |
|---|---|---|---|---|---|---|---|
| | | 2011-2014 | 2015-2017 | 2018-2020 | | | |
| Neurologic deterioration, % | IVT only (n=6447) | 17.8 | 13.8 | 13.7 | 15.3 | 0.001 | 0.003 |
| | EVT only (n=3076) | 22.4 | 13.4 | 14.4 | 15.7 | 0.001 | <.001 |

EVT indicates endovascular thrombectomy; IVT, intravenous thrombolysis.

*Adjusted for age, sex, initial NIHSS score

**eTable 6. Comparisons of Patient Characteristics between the CRCS-K-NIH registry and National Stroke Audit Program**

| | | Men, % | Age, mean (SD) | | Onset to arrival time, median hour (IQR) | NIHSS, median (IQR) |
|---|---|---|---|---|---|---|
| | | | Men | Women | | |
| 2013-2014 | CRCS-K | 58.1 | 64.8 (12.9) | 70.9 (12.7) | 5.7 (1.8-24.5) | 4 (1-8) |
| | NSADB | 57.7 | 66.1 (12.3) | 73 (11.8) | 6.3 (1.7-24.5) | 3 (1-7) |
| 2016 | CRCS-K | 58.3 | 65.4 (12.6) | 71.5 (12.7) | 5.3 (1.6-23.4) | 3 (1-8) |
| | NSADB | 57.7 | 66 (12.5) | 73.6 (12.1) | 5.7 (1.6-22.9) | 3 (1-8) |
| 2018 | CRCS-K | 57.8 | 65.9 (12.7) | 72.7 (12.7) | 5.7 (1.6-24.6) | 3 (1-7) |
| | NSADB | 57.9 | 66.9 (12.5) | 74.1 (12.5) | 6.2 (1.6-24.2) | 3 (1-7) |

IQR indicates interquartile range; NIHSS, National Institutes of Health Stroke Scale; NSADB, national stroke audit database; SD, standard deviation.
